# Group A streptococcal meningitis with the M1_UK_ variant in the Netherlands

**DOI:** 10.1101/2023.02.27.23286423

**Authors:** Boas C.L. van der Putten, Bart J.M. Vlaminckx, Brechje de Gier, Wieke Freudenburg-de Graaf, Nina M. van Sorge

**Affiliations:** National Institute for Public Health and the Environment, Bilthoven, Netherlands; Department of Medical Microbiology and Immunology, St Antonius Hospital, Nieuwegein, Netherlands; Department of Medical Microbiology and Infection Prevention, Amsterdam UMCs, Amsterdam, Netherlands; Netherlands Reference Laboratory for Bacterial Meningitis, Amsterdam UMCs, Amsterdam, Netherlands

## Abstract

We analyzed over 40 years of bacteriological surveillance data to reveal a 4-fold increase in Group A streptococcal (GAS) meningitis in 2022 compared to the annual average of 1982-2021 (n=5/year). Already 10 GAS meningitis cases occurred in 2023 (until March 13^th^). Molecular typing revealed that 25 out-of-29 (86%) isolates received in 2022 and 2023 were *emm*1.0 (Figure 1). WGS analysis of 19 *emm*1.0 isolates (2019 until 20^th^ December 2022) demonstrated that 15 out-of-19 (79%) isolates belonged to the toxicogenic M1_UK_ lineage. Based on these observations, we urge clinicians to be vigilant regarding clinical sign of meningitis with invasive GAS infections, since this disease manifestation appears to have a higher than expected occurrence due to clonal replacement by the recently-emerged M1_UK_ variant.

## Introduction

*Streptococcus pyogenes* can cause a wide range of invasive infections, collectively referred to as invasive group A streptococcal (iGAS) infections. A rare but severe disease manifestation of iGAS is meningitis, which has an estimated incidence in adults of 0.2 per 1,000,000 persons per year ^1,2^. Several European countries as well as the United States have observed an increase of iGAS infections in the past months, with shifts in affected age groups and clinical presentations relative to historic observations ^3,4^. Here we performed an epidemiological assessment of *S. pyogenes* meningitis cases, defined by culture-positive cerebrospinal fluid (CSF), using over 40 years of national bacteriological surveillance data.

## Methods

Active microbiological surveillance and molecular epidemiology of all-cause bacterial meningitis, including *S. pyogenes*, is performed by the Netherlands Reference Laboratory for Bacterial Meningitis (NRLBM). We analyzed *S. pyogenes* isolates cultured from CSF between 1982-2023 (until March 13^th^). Typing of *S. pyogenes* has been based on sequence variation in 180 bp of the *emm* gene and is routinely performed since 2013 according to the Centers of Disease Control and Prevention protocol (https://www.cdc.gov/streplab/groupa-strep/resources.html). To distinguish between the recently-identified toxicogenic M1_UK_ variant and the M1_global_ strains ^5^, *emm*1.0 isolates received between 2019 and 20^th^ Dec 2022 were analyzed by whole-genome sequencing (WGS; Illumina HiSeq). In accordance with Dutch law, approval by a medical ethics committee was not deemed necessary since cases were not subject to any actions or rules of conduct. No informed consent was requested and only pseudonymized data were used for the study.

## Results

Between 1982 and 2021, the NRLBM received 216 *S. pyogenes* CSF isolates, with an annual mean of five isolates (range 1-15). In contrast, 19 *S. pyogenes* CSF isolates were received in 2022, and 10 isolates were received between January 1 and March 13, 2023.

Among the 48 CSF isolates from 2013-2021, 19 different *emm* (sub)types were distinguished, with *emm*1.0 being the dominant *emm*-type at 35% (n=17; Figure 1)^6^. However, from the 29 *S. pyogenes* CSF isolates that have been received between 2022 and 13^th^ March 2023, 25 out-of-29 (86%) isolates were *emm*1.0 (Figure 1). Using WGS analysis for a smaller selection of *emm*1.0 isolates (2019 until 20^th^ December 2022), 15 out-of-19 (79%) isolates belonged to the toxicogenic M1_UK_ lineage^5^, with all *emm*1.0 isolates received from May 2022 onwards representing the M1_UK_ variant. The 15 M1_UK_ isolates originated from different municipalities, spread across the Netherlands. The age distribution of meningitis patients caused by *emm*1.0 strains was similar in 2022-2023 compared to 2013-2021 (Figure 2).

**Figure 1.**
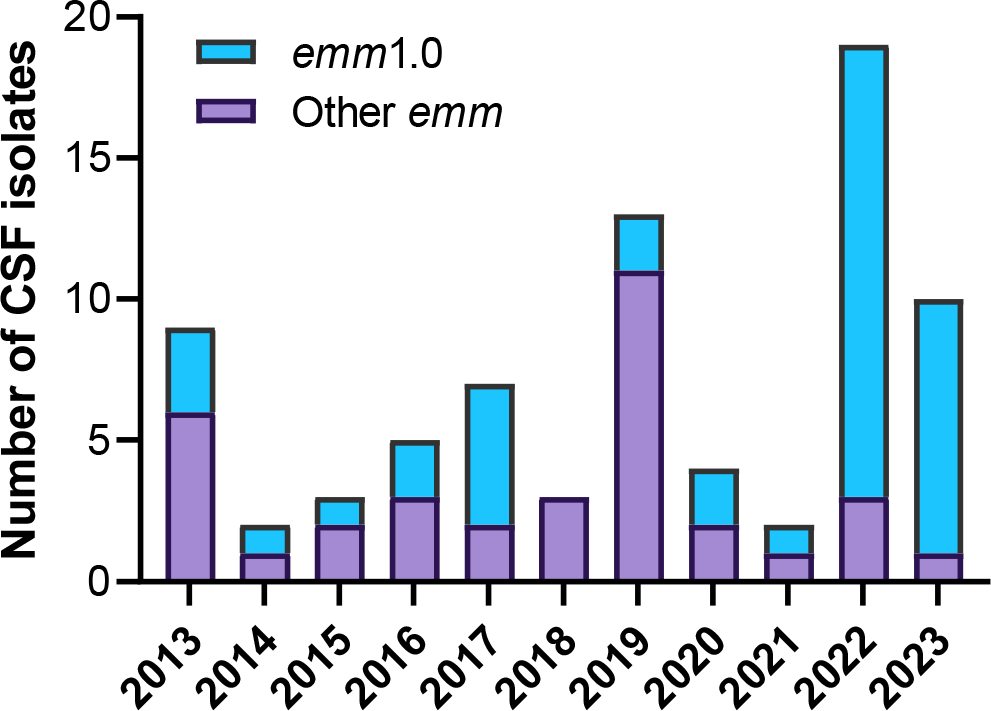
Number of received *S. pyogenes* CSF isolates with proportion of *emm*1.0, 2013-2023 (until March 13^th^).

**Figure 2.**
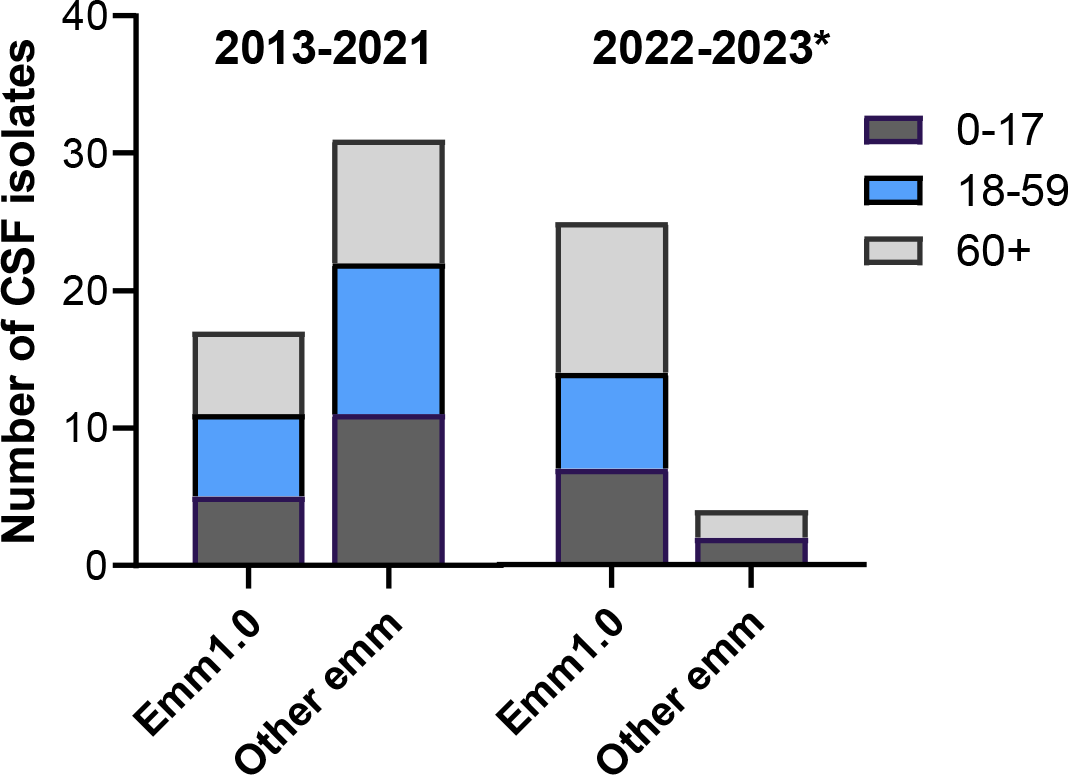
Age distribution of *S. pyogenes* meningitis patients infected with *emm*1.0 or other *emm* types in 2013-2021 versus 2022-2023. * Until March 13^th^ 2023.

## Discussion

Since 2022, an increase in *S. pyogenes* meningitis, with strong overrepresentation of the *emm*1.0 type and M1_UK_ variant, has been observed in the Netherlands. This study considered meningitis patients based on *S. pyogenes* cultured from CSF to provide a consistent comparison over time. Study limitations include the exclusion of meningitis patients with only a positive *S. pyogenes* blood culture, resulting in an underestimation of the overall national occurrence of *S. pyogenes* meningitis, and the small number of CSF isolates with limited patient information, which precluded more in-depth statistical and clinical evaluation. Furthermore, submission bias to the NRLBM may play a role, given the increased general submission of *S. pyogenes* isolates throughout 2022. Data from other countries are needed to assess whether the here observed increase in *S. pyogenes* meningitis is more widespread. The strong overrepresentation of *emm*1.0 among the received CSF isolates underlines the invasiveness of this *emm* type ^5^. Moreover, the dominance of the toxicogenic M1_UK_ variant among *emm*1.0 CSF isolates suggests an extensive replacement of circulating *S. pyogenes emm*1.0 strains. Detailed clinical studies are required to assess whether iGAS infections caused by the M1_UK_ variant are generally more severe. Clinicians are urged to be vigilant regarding the occurrence of meningitis with *S. pyogenes*, as this disease manifestation appears to have a higher than expected incidence with the recently emerged M1_UK_ variant.

## Data Availability

The mentioned strains are available upon request to the authors.

## Acknowledgements

The authors would like to thank Agaath Arends-van ‘t Klooster, Wendy Bril-Keijzers, Ilse de Beer-Schuurman, Jolein Pleijster, and Claudia Burger for their continued contributions as research technicians of the NRLBM (Amsterdam UMCs, Department of Medical Microbiology and Infection Prevention, Amsterdam, The Netherlands). Data are available upon request to the corresponding author with exception of specific patient information which will only be provided in aggregated form. NMvS declares fee for service and consultancy fees directly paid to the institution from MSD and GSK outside the submitted work. NMvS declared royalties related to a patent (WO 2013/020090 A3) on vaccine development against *Streptococcus pyogenes* (Vaxcyte; Licensee: University of California San Diego with NMvS as co-inventor). NMvS is a member of the Science advisory Board for the ItsME foundation (unpaid) and Rapua te me ngaro ka tau project (paid to institution; Project facilitating Strep A vaccine development for Aotearoa New Zealand). Other authors declare no conflict of interest.

## Notes

### Funding Statement

This study did not receive any funding

### Summary of Updates

Fixing/formatting of references

